# What percentage of severely impaired stroke survivors retain residual voluntary EMG?

**DOI:** 10.64898/2026.08.03.26359564

**Authors:** Monisha Yuvaraj, Steffi Graff, Prabhakar Appaswamy Thirumal, Sanjith Aaron, Ander Ramos-Murguialday, Nebojsa Malesevic, Christian Antfolk, Etienne Burdet, SKM Varadhan, Sivakumar Balasubramanian

## Abstract

**Back-ground:** Beneficial rehabilitation interventions for severely impaired stroke patients are limited. Owing to practical constraints in the routine clinical use of electroencephalogram (EEG)-based brain-computer interfaces, this study investigates the feasibility of using a more practical electromyography (EMG) to detect movement intention in severe stroke subjects without visible movement. Currently, no large-scale studies provide strong evidence in favour of EMG-based human-machine interaction for closed-loop control of robotic assistance for severe stroke.

**Objective:** To screen severely impaired stroke subjects without active wrist extension for the presence of residual EMG activity.

**Methods:** High-density surface EMG was recorded from the wrist extensor muscles of 100 severely impaired stroke survivors while they repeatedly attempted wrist extension. EMG activity during “Rest” and “Move” states was compared, and subjects showing statistically greater muscle activity during Move than Rest were classified as having residual EMG. A modified Hodges detector combined with the probability difference-sum ratio (PDSR) was used for classification, with a threshold of 0.73 identifying subjects with residual EMG.

**Results:** Of the 100 subjects without active wrist extension (Muscle power: MRC < 2), 64 exhibited residual EMG activity, supporting the feasibility of EMG for movement intention detection. Among these, 35 demonstrated consistent muscle activity (Detection probability > 0.2); representing suitable candidates for EMG-driven robot-assisted therapy.

**Conclusions:** A substantial proportion of severely impaired stroke subjects without active movement could benefit from a simpler EMG-based interface for robot-assisted therapy. Distinct neural mechanisms (intact voluntary drive or abnormal co-activation) may contribute to the residual muscle activity, which should be considered while designing control strategies.

## Introduction

Substantial recovery of sensory-motor function after stroke is possible with high-intensity, high-dose movement training^1^. However, recovery after stroke largely depends on the integrity of residual corticospinal tract (CST)^2^ which determine the extent and speed of recovery^3^. Approximately one-third of stroke survivors present with severe hemiplegia and complete loss of voluntary movement^3,4,5,6^. Because of their perceived poor recovery potential and the associated clinical challenges, rehabilitation research and therapeutic options for this population remain scarce^2^. Robot assisted therapy contingent on movement intention enables temporal coupling between the voluntary effort and afferent feedback from assisted movement, strengthening cortical connections through Hebbian plasticity^7^. However, detecting movement intention in severe stroke without residual movement is a challenge.

Brain–computer interfaces (BCIs) that decode intention from brain signals to control assistive devices or functional electrical stimulation (FES) are promising approaches to promote active engagement during training in severe stroke^8,9,10^. However, several limitations of electroencephalography (EEG)-based BCIs restrict their routine clinical application, including BCI illiteracy^11^, low signal-to noise ratio^12^, long calibration times due to substantial inter-subject variability^13,14^, and lack of task specificity^14^. In addition, the event related desynchronisation (ERD) may occur in anticipation of robot-imposed passive movements rather than reflecting voluntary motor intent ^15^. For widespread adoption or home use, BCIs need to be simple, robust, and easy to deploy, thus motivating the exploration of simpler alternatives such as electromyography (EMG)^16^. However, a key question is whether severely impaired stroke survivors without visible voluntary movements have sufficient residual EMG activity to enable movement intention detection.

DiPietro et al.^17^ demonstrated residual EMG activity in a stroke survivor despite the absence of observable movement. More recently, Steele et al. detected muscle activity in 11 out of 21 acute stroke survivors with a manual muscle test score of 0^18^. Since, EMG was monitored continuously during acute inpatient care, it remained unclear whether the detected activity was solely volitional^18^, highlighting the need for a well-designed experimental protocol to convincingly establish presence of detectable residual EMG in severe stroke.

Given this context, our previous work demonstrated that approximately 73% (22 out of 30) of severely affected stroke survivors without visible wrist or finger extension had residual voluntary EMG^16^. In a subsequent study, an automated approach to optimise the detector parameters increased this proportion to approximately 80%^19^. However, both findings were secondary analyses of data from a randomized controlled trial on BCI-based robot-assisted training^9^ with a limited sample size, which warrant further investigation.

To this end, the current work presents a large screening study designed to address the question of **what percentage of severely impaired stroke survivors with no visible voluntary movements retain residual, detectable voluntary surface EMG?** If this percentage is substantial, similar to our previous preliminary results^16^, this would support investing efforts towards developing EMG-based motor intent detection interfaces. EMG-based systems are a simpler alternative to actively engage severely impaired stroke participants in assisted therapy, which may have long-term clinical implications for this population who currently have little or no therapeutic options.

## Methods

### Participants

The study was approved by the Institutional Review Board of Christian Medical College (IRB Min. No. 2502130; dated 26.02.2025), Vellore, and conducted at the Department of Neurology, Christian Medical College Vellore, Ranipet, Tamil Nadu, India. Sample size was estimated using the single-proportion method based on our previous study, in which 22 of 30 subjects showed residual EMG (73%). Assuming the same prevalence, a 10% margin of error, and a 95% confidence level, the required sample size was 80; to allow for exclusions, 100 participants were recruited. Eligible participants were stroke survivors aged 18-75 years with upper-limb weakness who were unable to actively extend the wrist against gravity (as assessed by the Medical Research Council (MRC) scale < 2). Exclusion criteria included severe cognitive impairment, allergy to the electrode material, or pain in the affected upper limb interfering with the recording session. All subjects underwent an initial examination to confirm that they are clinically stable and suitable for participation in the study. Written informed consent was then obtained prior to starting with the experiment, after the study objectives and procedures had been clearly explained to them.

### Experimental Set-up and Protocol

EMG signals were recorded using a commercial 400-channel Quattrocento (OT Bioelettronica, Turin, Italy). Participants were seated with the hand attached to the experimental setup and the forearm secured in a neutral position on an armrest to minimize compensatory movements (Supplementary Figure S3). An optical encoder integrated into the setup monitored any angular deviation during movement attempts. Two 64-channel high-density surface EMG (HD-EMG) matrix electrodes (GR10MM0808, OT Bioelettronica; 8 × 8 grid, 10 mm inter-electrode distance) were placed over the dorsal and volar aspects of the forearm to record activity from the wrist extensor and flexor muscles, respectively. The grids were centred approximately one-third of the distance from the elbow to the wrist, aligned with the forearm axis, and positioned to maximize muscle-specific recordings while minimizing crosstalk. A detailed description of the recording procedure is provided in Supplementary Material.

Before the experiment, cognitive function and spasticity were assessed using the Montreal Cognitive Assessment (MoCA) and Modified Ashworth Scale (MAS), respectively. Wrist extensor muscle strength was then evaluated after securing the forearm in the armrest, using the Medical Research Council (MRC) scale to confirm participant’s eligibility. All assessments were conducted by a trained therapist.

Participants were instructed to alternately relax and attempt movement while surface EMG was recorded. Auditory and visual cues guided each trial. During the move phase, participants were instructed to attempt wrist flexion or extension in response to a “flex” or “extend” cue. Each trial consisted of a 10-s rest phase, a 3-s “ready” cue to minimize anticipatory movement in the rest period, and a 5-s move phase. (Figure 1, right). Ten trials constituted one block, and each participant completed 10 alternating blocks of wrist flexion and extension attempts (five blocks each, 50 trials per movement type). A few minutes of rest was provided between blocks, as needed, to minimize fatigue.

**Figure 1:**
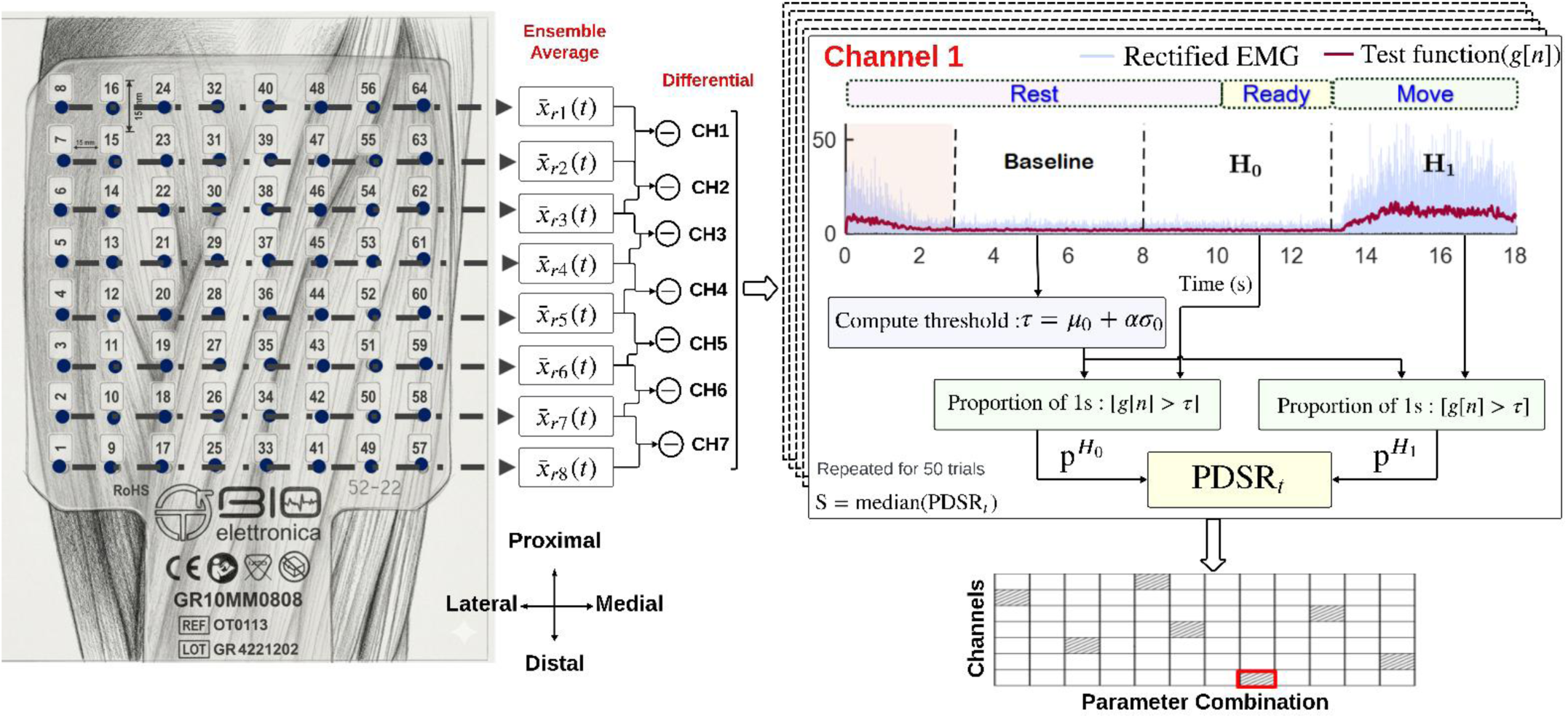
Schematic illustration of the best channel selection pipeline. **Left:** Conversion of the 64-channel HD-EMG recordings into seven differential channels (Generated using Google Gemini (Google DeepMind) and reviewed by the authors for accuracy), where 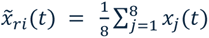 is the ensemble average of EMG recordings across the electrodes in the *i^t^*^ℎ^ column. Differential channels are then obtained by subtracting successive column averages, 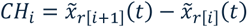. **Right:** Pipeline for identifying the *best channel* for each participant, which is used to determine the presence or absence of residual EMG activity. For each channel, the detector parameters yielding the maximum PDSR are identified (grey-shaded box in the grid). The channel with the highest PDSR across all channels is selected as the best channel. The illustrated example shows the 7th channel being identified as the optimal channel for classification.

### Data Analysis

EMG signals were recorded in a monopolar configuration at a sampling frequency (*fs*) of 2048 Hz. The raw signals were filtered using a 2nd-order Butterworth high-pass filter (10 Hz cut-off) and an IIR comb filter to suppress 50 Hz power-line interference and its harmonics. Channels exhibiting excessive amplitude variation or poor signal quality (“bad channels”) were excluded from further analysis. The definition of bad channel and a detailed procedure to remove it is explained in the supplementary material (Appendix B). As the objective was to detect the presence of activity in any of the muscles within the wrist extensors and flexors, the 64-channel HD-EMG recordings were transformed into seven representative bipolar channels. This was achieved by ensemble averaging channels across columns (orthogonal to the muscle fibre direction) and computing the difference between adjacent column averages (Figure 1).

To estimate the percentage of severely impaired stroke survivors with detectable residual EMG, we compared EMG recordings under two experimental conditions, as described in our previous work^19^:

- <u>Null hypothesis</u> (*H*_0_)∶ When the subject is relaxed, the EMG recording contains only baseline activity.
- <u>Alternative hypothesis</u> (*H*_1_): When the subject attempts a movement, the recordings may contain intermittent muscle activity in addition to baseline activity.

A subject was classified as having residual EMG if the EMG activity observed under move phase (*H*_1_) exceeds that observed under rest phase (*H*_0_) condition.

Let *x_T_*(*t*) denote the time series of the recorded EMG signal for a given trial T. The first 3 s of each trial (red-shaded region in Figure 1: right side) were discarded to avoid persistent muscle activity from the previous trial. The remaining 15 s were divided into three consecutive 5-s segments. The first segment (Baseline: 3–8 s) was used to estimate mean (*μ*_0_) and standard deviation (*σ*_0_) required for computing the threshold (*τ*) to determine the presence or absence of muscle activity. The remaining two segments are used to construct the *H*_0_ and *H*_1_ datasets as follows:

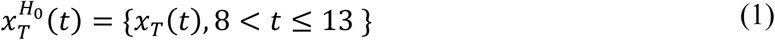

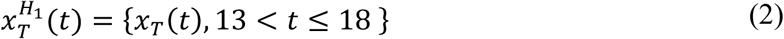

Involuntary EMG spikes observed in the data, contaminate voluntary EMG activity and adversely affect classification performance^20^. Therefore, trials containing these spurious spikes were first identified as trials exhibiting high dissimilarity between the actual baseline data and the synthetic baseline data (generated using an AR model). These trials were then filtered using the wavelet denoising approach as described in our previous work ^19^. The trials were split into the *H*_0_ and *H*_1_ segments after the removal of the involuntary spikes.

Muscle activity was detected using a modified Hodges detector, which consistently showed superior performance in our previous studies^19,21^. The detector maps the processed EMG signal into a binary signal, assigning a value of 1 when muscle activity is detected and 0 otherwise. The proportion of 1s (detection probabilities: 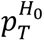 and 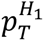) in the *H*_0_ and *H*_1_ segments was compared to quantify the difference in the amount of muscle activity in the two segments. Detector parameters were optimized using a grid search to maximize the separation between *H*_0_ and *H*_1_ conditions. This separation was quantified using the probability difference sum ratio (PDSR) metric which was shown to have superior performance compared to other separation measures in our previous work^19^. The modified Hodges detector optimized using the PDSR achieved low detection latency and high classification accuracy^19^. The PDSR measure also was found to be an effective screening measure for identifying subjects with residual EMG. Therefore, the same PDSR-based optimization and classification approach was adopted in the present study. The current study performed a channel-specific optimization of the modified Hodges detector using PDSR across the seven bipolar channels. The channel with the highest PDSR was selected as the *best* channel for assessing the presence or absence of residual EMG activity. This procedure was performed separately for the flexor and extensor electrode grids during both flexion and extension attempts. However, since wrist extension deficits are more common after stroke and flexor muscles typically recover earlier than the extensors^22,23^ the present analysis focused on residual EMG activity in the wrist extensor muscles during wrist extension attempts. Therefore, participants exhibiting no visible wrist extension movement were recruited even if they had active wrist flexion.

### Statistical Analysis

To cross-verify whether the increase in detection probability from rest to move 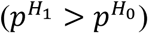 differed between subjects with and without residual EMG, a linear mixed-effects model (LME) was fitted with condition (*H*_0_, *H*_1_) and group (with/without-residual EMG) as fixed effects, along with their interaction term (condition × group). If a significant interaction was observed, the simple effects of condition were examined using independent group-specific LME models, with the effect size quantified as the estimated marginal mean difference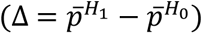.

Within-subject differences between the distributions of 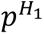 and 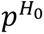 were compared using the Wilcoxon signed-rank test (1% significance). The secondary analysis of the effect of clinical assessments and demographic characteristics on the presence or absence of residual EMG was investigated by comparing the PDSR values across subgroups using the Wilcoxon rank-sum test. The effect of time since stroke was evaluated using the Kruskal–Wallis test. All signal processing and statistical analyses were performed in MATLAB R2023b (MathWorks, Natick, MA, USA).

## Results

Participants were recruited as outpatients from the Department of Neurology (stroke clinic), CMC Vellore, between February 2025 and May 2026. Of approximately 350 stroke survivors screened for eligibility, those with severe contractures preventing attachment to the experimental setup, inability to follow instructions, or the ones with comorbidities (severe cough, fatigue, or emotional instability) affecting the recording session were excluded (Supplementary Figure S4). Six participants who did not complete the recording session were also excluded from the analysis. Demographic and clinical characteristics of the recruited 100 stroke survivors are summarised in Table 1. Clinical assessments were conducted before the EMG recording session; ensuring assessor blinding to the primary outcome (presence or absence of residual EMG). MAS scores were available only for 71 subjects. In addition, due to a technical error, the angle data were not recorded for 5 subjects, though their EMG recordings could still be used for the analysis.

**Table 1:** Characteristics of the study’s stroke participants (n = 100)

| **Demographics** |  |
| --- | --- |
| Age, years - mean (SD) | 46.4 (11.6) |
| Sex - male/female | 78 / 22 |
| Type - ischaemic/ Haemorrhagic* | 57 / 26 |
| Affected side - left / right | 56 / 44 |
| Time since stroke |  |
| Early subacute (< 3 months) | 6 |
| Late subacute (3-6 months) | 7 |
| Chronic (> 6 months) | 77 |
| **Clinical measures** |  |
| MRC grade - 0 / 1 | 51 / 49 |
| MAS flexor tone - median (IQR)<sup>a</sup> | 2 (3) |
| MAS extensor tone - median (IQR)<sup>a</sup> | 0 (0) |
| MoCA - mean (SD) | 16.6 (7.3) |
| *The stroke etiology of 17 subjects was not known; <sup>a</sup> MAS data were recorded only for 71 participants since the protocol was amended later in the study. MAS = Modified Ashworth Scale; MoCA = Montreal Cognitive Assessment; MRC = Medical Research Council; SD = standard deviation; IQR = interquartile range. |  |

### Percentage of severely impaired stroke survivors with residual extensor EMG

The EMG recording from the identified *best* channel, that exhibits maximum separation between the *H*_0_ and *H*_1_ was used to determine the presence or absence of residual EMG activity. Participants with a PDSR exceeding the predefined threshold of 0.73 were classified as having residual wrist/finger extensor EMG. This threshold was determined from the observed distribution of the PDSR values, similar to our previous work^19^. The kernel density estimate(KDE) of the PDSR showed a dip between 0.7-0.8 (Figure 2a), and a threshold of 0.73 was selected (dashed red line in Figure 2a), corresponding to the lower bound of the 95% bootstrap confidence interval for the KDE-derived dip location (0.769, 95% CI: 0.730–0.809; Supplementary Appendix E). Overall, **64% (64/100) of participants exceeded the threshold of 0.73 for the PDSR and were classified as having residual EMG activity**. The detection probability remained close to zero below this PDSR threshold and increased sharply beyond it. Figure 2a also shows a moderate positive correlation between the PDSR separation measure and the detection probability under *H*_1_ (Spearman’s *r* = 0.79, *p* < 0.001) indicating that larger separation measures were associated with higher detection probabilities.

**Figure 2.**
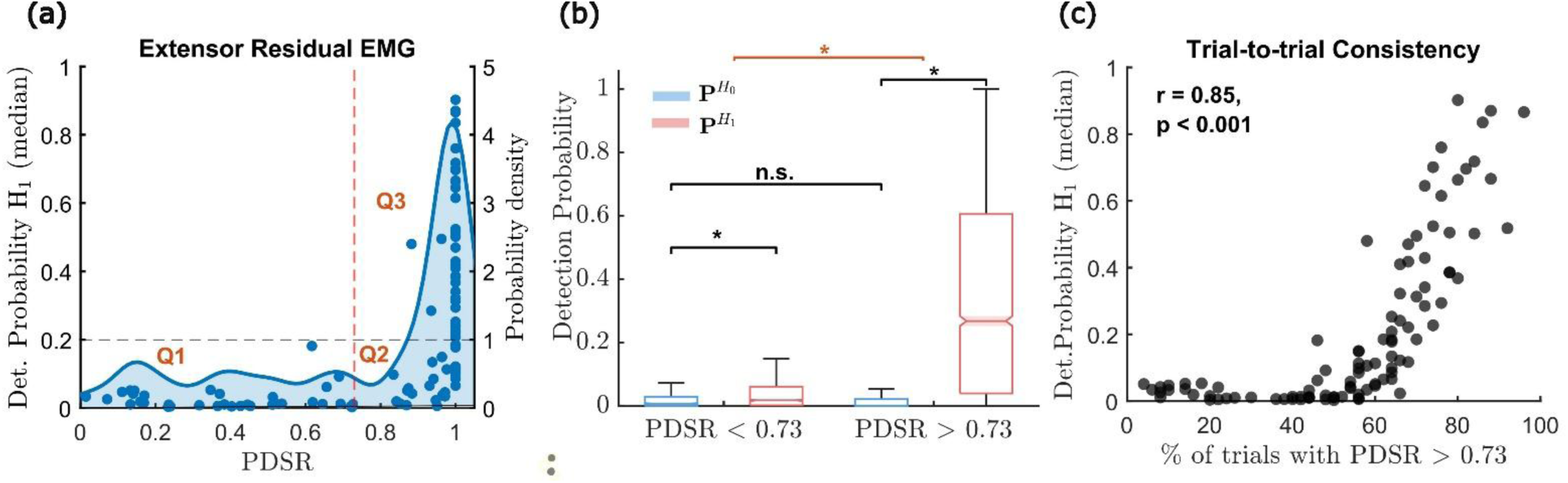
Presence/Absence of residual EMG: (a) The scatter plot of the optimum separation measure – PDSR and the median detection probability computed with EMG recorded during extension attempts. The kernel density plot of the PDSR separation measure is shown as the filled blue trace, with the PDSR threshold of 0.73 (dotted red line) to identify subjects with residual EMG. The horizontal dashed line is drawn at detection probably of 0.2. (b) Boxplot of detection probability under the *H*_0_ and *H*_1_ of all trials for subjects with and without residual EMG (outliers ignored for plotting). * indicates statistical significance with *p* < 0.001 and the red line corresponds to a significant interaction effect between the two groups. (c) The scatter plot of the percentage of trials with PDSR > 0.73 and the detection probability under *H*_1_ (median) to indicate consistency across trials. For a detection probability of greater than 0.2, more than ≈ 60% trials have good separation between the EMG activities in their respective rest and move phases.

#### Differences between 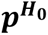 and 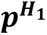 in participants with and without residual EMG

A linear mixed effect model was fitted to understand the differences in 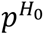 and 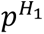 between the participants with (PDSR > 0.73) and without residual EMG. Figure 2b presents a summary plot of 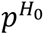 and 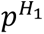 for all 100 participants, separated into two groups based on the presence/absence of EMG. This plot immediately reveals that 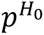 was the same in both groups (difference is not significant). On the other hand, 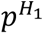 was significantly higher in the EMG group compared to the no EMG group, which explains the higher PDSR in this group. The linear mixed-effects model revealed a significant condition × group interaction effect (F (1, 100.06) = 66.11, p<0.001), indicating that the difference between 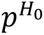 and 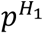 is significantly different between subjects with and without residual EMG. The difference in mean 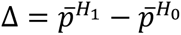 was substantially larger in the group with residual EMG (Δ = 0.305, p < 0.001) than in the group without residual EMG (Δ = 0.034, p<0.001). Furthermore, a Wilcoxon signed-rank test for individual participants comparing 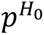 and 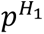 showed significantly higher 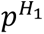 than 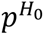 for all participants classified as having residual EMG (Supplementary Table S1). Thus, supporting the conclusion that 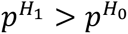 in participants deemed to have residual EMG.

#### Trial-to-trial consistency of residual voluntary EMG generation

The consistency of residual EMG generation by individual participants was quantified as the percentage of trials with PDSR > 0.73. Figure 2c shows a strong positive correlation between trial consistency and median detection probability under H_1_ (Spearman’s r = 0.85; p < 0:001), indicating that participants with higher 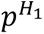 generally show more trials with better separation between the rest and move phase EMG activity. Figure 2c also reveals that participants with 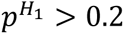 produce ≈ 60% or more trials with good separation in EMG activities between the rest and move phases.

### Relevance of clinical measures on the presence of residual EMG

#### Does extensor muscle power correlate with residual muscle activity?

The study only included subjects with MRC grade less than 2; MRC grade 0 indicates no visible/palpable contraction, and MRC grade 1 indicates flicker or trace of contraction without observable joint movement. Therefore, it is reasonable to expect that subjects with MRC grade 1 are more likely to exhibit residual EMG activity than those with MRC grade 0. Figure 3a presents box plots of the PDSR separation measure for the MRC 0 (n = 51) and MRC 1 (n = 49) subgroups. As expected, most subjects with MRC grade 1 exhibited PDSR values above the classification threshold 0.73 and had significantly higher separation measures than subjects with an MRC grade 0 (p < 0.001).

**Figure 3.**
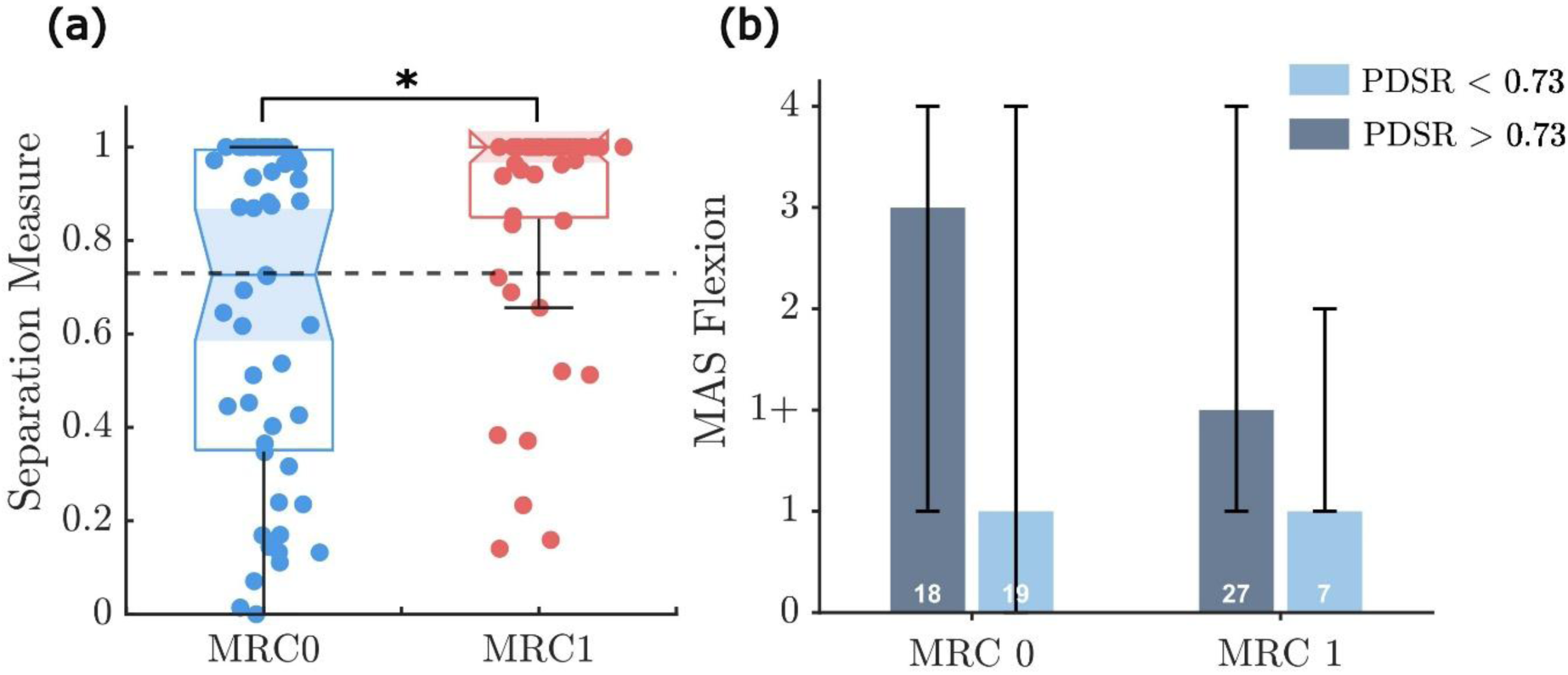
Relevance of clinical measure on presence/absence of residual EMG: (a) Stratification based on the muscle power (MRC grade): Boxplot of the separation measure of subjects with MRC 0 (N = 51) and MRC 1 (N=49). Subjects with MRC 1 show good separation from those with MRC 0 (p < 0.001). (b) Bar graph of median MAS in the flexors for subjects with and without residual EMG for MRC grade 0 and 1 subgroups. The IQR is represented as an error bar. The data of only the 71 subjects whose MAS scores are recorded are presented in the bar graph.

#### Does spasticity contribute to the residual muscle activity?

Figure3a shows that some participants (n = 18) with an MRC-0 were classified as having residual EMG activity. To explore the possible explanations for this observation, we examined the median MAS score for the flexor muscles (assessed by passive extension of the wrist) for subjects with and without residual EMG activity within the MRC 0 and MRC 1 subgroups separately. In both subgroups, subjects classified as having residual EMG tended to have higher MAS scores than those classified as not having residual EMG (Figure 3b), although this trend was not significant (MRC 0: p=0.0655; MRC 1: p=0.0880). All subjects with MRC 0 and MAS 0 were classified as not having residual EMG, suggesting that detectable residual muscle activity is unlikely in a completely flaccid hand. Furthermore, none of the subjects with an MRC 1 had an MAS score of 0 (Figure 3b), indicating a connection between the presence of voluntary muscle activation and increased muscle tone probably due to spasticity in this cohort.

##### Effect of participants’ demographic characteristics on the presence of residual EMG

To better understand the difference between the participants with and without residual EMG, we investigated the association between the PDSR value for each individual with various demographics variables: time since stroke, affected side, stroke aetiology, gender and age. Figure 4a presents boxplots of the PDSR separation measure for the three post-stroke phases: early subacute, late subacute, and chronic^24^. The Kruskal-Walli’s test revealed a significant effect of time since stroke on the separation measure (H (2) =6.13, p=0.047). Participants in the early subacute phase (< 3months) predominantly exhibited a separation measure below 0.73; post hoc analysis showed significantly lower values than the chronic group (p=0.041), suggesting that residual EMG activity may become more prominent with increasing time since stroke. However, the late subacute group (3–6 months post-stroke) did not differ significantly from either the early subacute (p=0.177) or chronic groups (p=1.00). Furthermore, no significant effect of stroke aetiology (p=0.1635), gender (p=0.122), affected side (p=0.276) or age (Pearson correlation: r=-0.0812; p=0.4217, Supplementary Figure S8) on the presence of residual EMG was observed.

**Figure 4.**
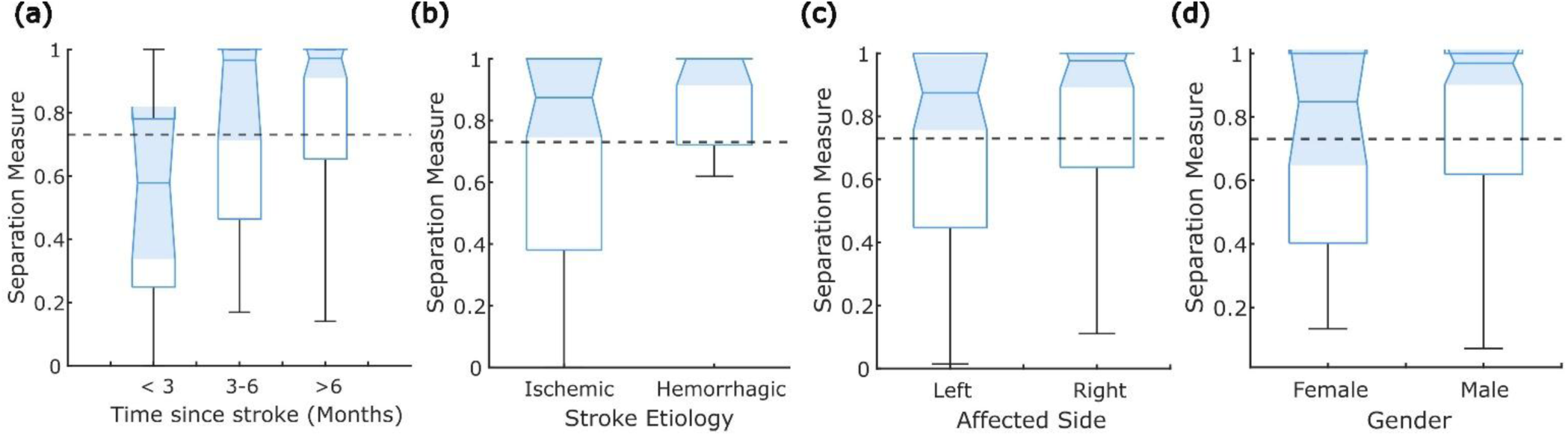
(a) **Effect of phase of stroke:** Boxplot of the separation measure (median PDSR) for subgroups divided based on time since stroke (Early sub-acute, late sub-acute, and chronic); (b) **Effect of stroke aetiology:** Boxplot of the separation measure (median PDSR) for ischemic and haemorrhagic stroke subgroups. (c) **Effect of affected side:** Boxplot of the separation measure (median PDSR) of left and right impaired stroke subjects. (d) **Effect of gender:** Boxplot of the separation measure (median PDSR) for subgroups divided based on gender. Black dotted line: The PDSR threshold for deciding presence/absence of residual EMG.

## Discussion

Effective rehabilitation interventions for severely impaired stroke survivors without visible movements remain limited. BCIs have emerged as promising rehabilitation tool for this population, however EEG-BCIs present practical barriers to routine clinical use. EMG could serve as a viable alternative to EEG-BCIs for detecting movement intention in severely impaired stroke survivors without observable movement. While preliminary evidence reported residual EMG in this population^16, 17,18,19^ there was no large-scale study in the literature providing strong evidence for the proportion of severe stroke survivors with residual EMG. The present study addresses this gap showing that 64% (64/100) of the stroke subjects without active wrist extension had detectable residual EMG, offering a strong empirical support for the clinical applicability of EMG-based intention detection for assisted hand neurorehabilitation in severe stroke. Although this percentage falls within the 10% margin of error assumed during sample size calculation, it is slightly lower than the 73% and 80% reported in our previous studies^16,19^. Three possible reasons for this difference could be:

a) Unlike our previous study^19^, where the *H*_0_ condition was simulated using synthetic baseline data generated using an autoregressive model²⁴, the current study derived *H*_0_ segments directly from the recorded EMG. The use of real baseline data likely reduced overestimation (0.8) by the PDSR separation measure (PDSR).
b) The current cohort included an equal number of MRC 0 and MRC 1 participants, with most subjects exhibiting very low PDSR values belonging to the MRC 0 subgroup. We do not have the MRC values for the previous cohort^9^. However, this cohort might have been predominantly MRC 1, since the PDSR values were all higher than 0.4 for all participants^19^. Furthermore, the present study also included participants in subacute stage of stroke, whereas the previous study exclusively included only chronic stroke survivors, which may have also contributed to the observed difference.
c) Posture effect over EMG activation ability: In the current work we are focusing on wrist extension and in our previous work the movements analysed were different and other muscle groups or activity patterns were involved. We know from previous work that residual upper arm motor function primes recruitment of paralyzed forearm muscles in stroke subjects^25^. Therefore, the arm configuration or posture in which the movement was executed conditions the ability to recruit the muscles involved in the movement.

Unlike the previous study, which optimized a single modified Hodges detector across two EMG channels²⁴, the current study has more spatial information as 64-channel HD-EMG recordings is utilised. Channel-specific detector optimization and systematic bad-channel removal, ensures the detection of weak muscle activity across the extensor muscles, making the results more reliable than our previous work.

A positive correlation was observed between the detection probability under *H*_1_ and the separation measure (PDSR). However, several subjects with very high PDSR values (> 0.73) still exhibit lower detection probability, implying that these subjects have more muscle activity when attempting movements than at rest, but not consistent enough to sustain the muscle activity for 5-s. This represents a major limitation of the separation measure as a standalone criterion for identifying subjects feasible for the EMG-based intent detection approach which can be addressed by employing an additional threshold of 0.2 for the median detection probability. As shown in Figure 2c, a detection probability ≥ 0.2 was generally associated with EMG activity being detected in more than 60% of trials, supporting the choice of this threshold. Figure 2a illustrates three regions divided by the thresholds of PDSR = 0.73 and median detection probability = 0.2. Based on these thresholds, we propose different control strategies for the subjects in each region. Approximately one third of the cohort (Figure 2a: Q1, *n* = 36) exhibited no detectable residual muscle activity and therefore cannot rely on EMG-based movement-intent detection, leaving EEG-based BCIs as the only alternative^26,27^. The remaining two-thirds (Figure 2a: Q2 and Q3, *n* = 64) can employ EMG to detect intention however the distinction lies in the control strategies. Subjects in Q2 region (*n* = 29) achieved a PDSR above the threshold but had a detection probability below 0.2, implying that these participants either produce intermittent bursts of muscle activity or cannot substance muscle activity over time; in such participants, the use of triggered robotic assistance^17,28^ may be a more appropriate approach. On the other hand, participants in the Q3 region (*n* = 35), exhibit sufficiently consistent and sustained EMG activity necessary for either on/off or continuous EMG-based control of robotic assistance^29,30^. Furthermore, subjects may transition between regions during rehabilitation, progressing from Q1 to Q2 as residual EMG activity emerges and from Q2 to Q3 as muscle activation becomes more consistent. Thus, a flexible control approach for robotic assistance may be warranted in such cases.

All participants recruited in the study could not actively extend their wrists. Kamper et al. reported that impaired finger extension following stroke result from of reduced voluntary drive to the agonist muscles, compounded by involuntary antagonist muscles activation due to spasticity and inappropriate co-contraction ^31^. These neurological mechanisms were reflected in our findings (Figure 3), where two subjects with similar muscle power (MRC = 0) differed in their EMG activity: one exhibited residual muscle activity, potentially attributable to abnormal co-activation (descending motor signals are present, but the “commands” are functionally inappropriate)^32,33^, whereas the other showed no detectable muscle activity, possibly reflection insufficient cortico-muscular control/drive of the recorded muscle (Supplementary Appendix F).

Severe motor impairment after stroke is associated with extensive CST damage^34,35^, which is compensated by the upregulation of alternative brainstem pathways, i.e., the reticulospinal tract (RST), and severe stroke survivors rely on the RST for any residual motor output^35^. Because, the RST primarily facilitates proximal muscle strength ^36^ and predominantly supports movements within abnormal flexor synergies, while limiting isolated and out-of-synergy motor control^37,36^ many participants recruited proximal muscles, i.e., executed shoulder abduction while attempting wrist movements. The preferential facilitation of flexor muscles by the RST, likely explains why many subjects exhibited wrist flexion during attempted wrist extension (Supplementary Figure S6).

Future work should investigate the neural mechanisms underlying residual muscle activity observed in our cohort and develop some data acquisition protocol including postural facilitation^25^. Furthermore, distinguishing between residual EMG arising from preserved voluntary corticospinal drive from that mediated by abnormal brainstem pathways could identify distinct patient subgroups and enable more targeted rehabilitation strategies. The result from this study must be interpreted in light of some of its limitations:

1. Although the subject’s hand was comfortably placed in a wrist flexion-extension module, trunk and shoulder movements could not be completely prevented. Therefore, the recorded EMG activity may partly reflect abnormal synergy patterns rather than solely voluntary wrist movement.
2. The proposed method may be influenced by experimental factors, such as the number of bad channels removed during preprocessing, variations in electrode placement, and the participant’s physiological state (e.g., fatigue following therapy), which could affect EMG recordings.

Based on the findings of the current study, we emphasise that EMG is a viable tool for movement intent detection in a subset of severely impaired stroke subjects with no visible movement. The observation that 64% of stroke survivors without active wrist extension exhibited detectable residual extensor EMG provides strong evidence supporting the feasibility of using EMG as a control signal for rehabilitation in this population. However, only 35% of these subjects demonstrated consistent muscle activity characterised by both good separation (PDSR > 0.73) and sufficient detection probability (> 0.2). This subgroup is therefore the most likely to be suitable for EMG-driven robot-assisted therapy. The present study also highlights several challenges associated with implementing EMG-driven robot-assisted therapy in individuals with severe stroke. These include practical difficulties in positioning the hand due to severe contracture, signal artifacts arising from involuntary muscle activity, and the complex neural mechanisms underlying residual EMG generation, all of which complicate reliable movement intent decoding. Physiological factors such as abnormal synergies, maladaptive and involuntary muscle activity should be carefully considered when designing control strategies for the severe stroke population. The study also highlights the heterogeneity among severe stroke population, suggesting that a single control strategy is unlikely to be optimal for all individuals. Future studies should evaluate the clinical effectiveness of subject-specific EMG-driven robot-assisted therapies. Longitudinal studies examining changes in muscle activity patterns across rehabilitation sessions, in terms of increases in amplitude and reductions in co-contraction and maladaptive muscle activity patterns, should be explored to predict recovery patterns and propose new EMG-based rehabilitation alternatives in severely impaired stroke survivors.

## Conclusions

The current study evaluated whether EMG could serve as an alternative to EEG-BCI for decoding movement intention in severely impaired stroke survivors without active wrist extension. This was done by analysing HD-EMG recordings from 100 stroke survivors with wrist extensor MRC of less than 2. Overall, the findings provide strong empirical support for the clinical applicability of EMG-based movement intent detection, with 64% of severely impaired stroke survivors exhibiting residual EMG activity in their wrist extensor muscles and 35% of them having sustained muscle activity during the move period (>20%). Additionally, we also highlight a systematic approach in screening severe stroke and identifying suitable candidates for EMG-driven robot-assisted therapy. Careful consideration of physiological factors and adopting a mechanistic approach to designing subject-specific control strategies may enhance the effectiveness of EMG-driven rehabilitation interventions.

## Data Availability

All data produced in the present study are available upon reasonable request to the authors

## Declaration of Conflicting Interests

The author(s) declared no potential conflicts of interest with respect to the research, authorship, and/or publication of this article.

## Funding

The author(s) disclosed receipt of the following financial support for the research, authorship and/or publication of this article: This work was supported by the Fluid Research Grant from CMC Vellore (grant number: IRB Min. No. 2502130; dated 26.02.2025).

## Notes

### Competing Interest Statement

The authors have declared no competing interest.

### Author Declarations

The study was approved by the Institutional Review Board of Christian Medical College (IRB Min. No. 2502130; dated 26.02.2025), Vellore

